# Estimating the impact of single-cell RNA sequencing of human tissues on drug target validation

**DOI:** 10.1101/2024.04.04.24305313

**Authors:** Emma Dann, Erin Teeple, Rasa Elmentaite, Kerstin B Meyer, Giorgio Gaglia, Frank Nestle, Virginia Savova, Emanuele de Rinaldis, Sarah A Teichmann

## Abstract

Whilst the use of single-cell RNA sequencing (scRNA-seq) to understand target biology is well established, its predictive role in increasing the clinical success of therapeutic targets remains underexplored. Inspired by previous work on an association between genetic evidence and clinical success, we used retrospective analysis of known drug target genes to identify potential predictors of target clinical success from scRNA-seq data. We investigated whether successful drug targets are associated with cell type specific expression in a disease-relevant tissue (cell type specificity), and with cell type specific over-expression in disease patients compared to healthy controls (disease cell specificity). Analysing scRNA-seq data across diseases and tissues, we found that both cell type and disease cell specificity are features enriched in targets entering clinical development, and that cell type specificity in the disease-relevant tissue is robustly predictive of target progression from Phase I to II. While scRNA-seq analysis identifies a larger and complementary target space to that of direct genetic evidence, its association with specificity and drug approval appears less clear. We discuss how further expansion and harmonization of single-cell datasets, more sophisticated integration of this data in target discovery, and improved methods for tracking clinical trial outcomes could enhance our ability to leverage scRNA-seq insights in drug development in future.

## Introduction

Drug discovery begins with the identification of candidate targets, drug-binding molecules whose modulation is hypothesized to be useful for the treatment of disease [1]. The discovery and development of a novel drug for a candidate target progresses in steps, from target validation to safety and efficacy clinical trials, and, in successful cases, regulatory approval. Development of a single new drug takes an average of 12-15 years and costs (including concurrent program failures) are estimated to range from 900 million – 2.6 billion USD per success [2,3]. A drug discovery program can fail at each step between early research to regulatory approval, and it is estimated that in >90% of cases failures can be attributed to suboptimal target selection for a given disease, resulting in safety or efficacy issues [4]. Together, these observations point to the need to improve the strategies and the data used in early stages of drug discovery to support the selection of candidate therapeutic targets, to increase the likelihood of clinical success.

Single-cell RNA sequencing (scRNA-seq) data is a particularly promising source of evidence for target selection, providing cell-level resolution of molecular profiles in disease-relevant tissues. Single cell technologies have already been applied extensively to characterize disease biology, in emerging diseases like COVID-19 [5,6], cancer [7–10], and common complex diseases across tissues [11–14]. The rapidly growing body of disease-relevant scRNA-seq data has already begun to inform the development of novel diagnostics and cell-targeting precision therapies [15]. This led us to ask to what extent information on cell type specific expression can boost the selection of promising drug targets.

Retrospective analysis of known drug targets has been used to identify features predictive of target success. Notably, such analyses have shown that targets linked to genetic variants associated with the relevant disease are twice as likely to reach clinical approval as targets with no genetic support [16–18]. These studies greatly impacted decision-making in biotech and pharmaceutical industries. Out of 428 newly FDA-approved drugs from 2013 to 2022, 271 (63%) are backed by direct or indirect human genetic evidence [19,20]. Even though establishing whether this influenced their discovery or development phases is difficult, 250 out of 271 genetics-backed drugs had publicly accessible genetic support before approval.

Given this precedent, here we used retrospective analysis to investigate whether common scRNA-seq disease analyses are informative as predictors of target clinical success.

## Results

Although scRNA-seq data can reveal several gene features useful for drug targeting, we focused on testing two features which are commonly derived from scRNA-seq studies and used as a way to validate genes of potential use as therapeutic targets: (1) *cell type specificity* in healthy disease-relevant tissue and (2) *disease cell specificity*, here referred to as the over expression in a cell type in a diseased compared to a healthy tissue (*Figure 1*). The reasoning behind highlighting these features is that drugs targeting *cell type specific* genes inhibit expansion and function of normal cells acquiring aberrant phenotypes in disease. For example, the GLP-1 receptor, targeted by commonly used anti-diabetic drugs, is normally expressed in pancreatic beta cells, which become dysfunctional in disease [13]. Conversely, drugs targeting *disease cell specific* genes suppress aberrant gene programmes directly. For example, inflammatory bowel disease patients are treated with antibodies targeting the tumor necrosis factor (TNF), which is known to be produced by various immune cell types and has been recently shown to be over-expressed in regulatory T cells, as well as other immune subtypes, in disease [21].

**Figure 1:**
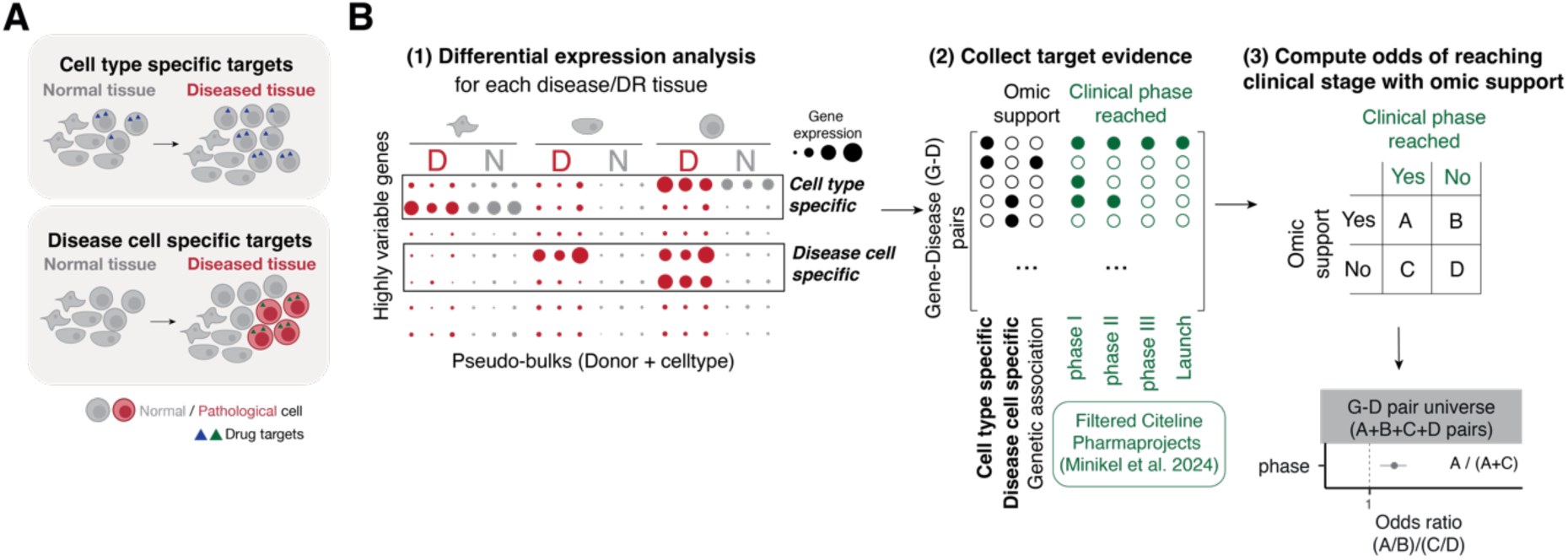
Overview of study design. (A) Illustration of rationale behind scRNA-seq support classes for target discovery: cell types expanding or acquiring aberrant function in disease can be targeted using cell type specific targets. Cells specifically expressing aberrant gene programmes can be targeted with disease cell specific targets (B) Workflow for analysis of association between scRNA-seq support and clinical success. We identify cell type specific and disease cell specific gene-disease pairs through differential expression analysis on pseudo-bulked data from the disease-relevant tissue (1). Data on genetic association and clinical success of targets was collected from the Citeline Pharmaprojects database, as processed by Minikel et al. 2024 (2). For each omic support class, we compute the odds ratio for the association between clinical success (passing clinical trials) and different classes of omic support (3).

We first considered diseases for which scRNA-seq data was available via the CZ CellxGene Discover database [22], defining a disease-relevant (DR) tissue for each disease term. Of the 58 disease terms in the CZ CellxGene database, 25 terms were retained for association analysis, based on availability of data from disease-relevant tissue and overlap with disease annotation terms in clinical data (see Supplementary Table 1 for a complete list of considered diseases and reasons to exclude from analysis). The most prevalent diseases were lung and immune disorders (Figure 2A). For each disease term, we collected gene expression count matrices and coarse cell type labels, harmonized using the Cell Ontology [23] (Figure 2B, Supplementary Figure 1, see Methods), for disease-relevant tissue samples from healthy and diseased individuals (Supplementary Table 2).

**Figure 2:**
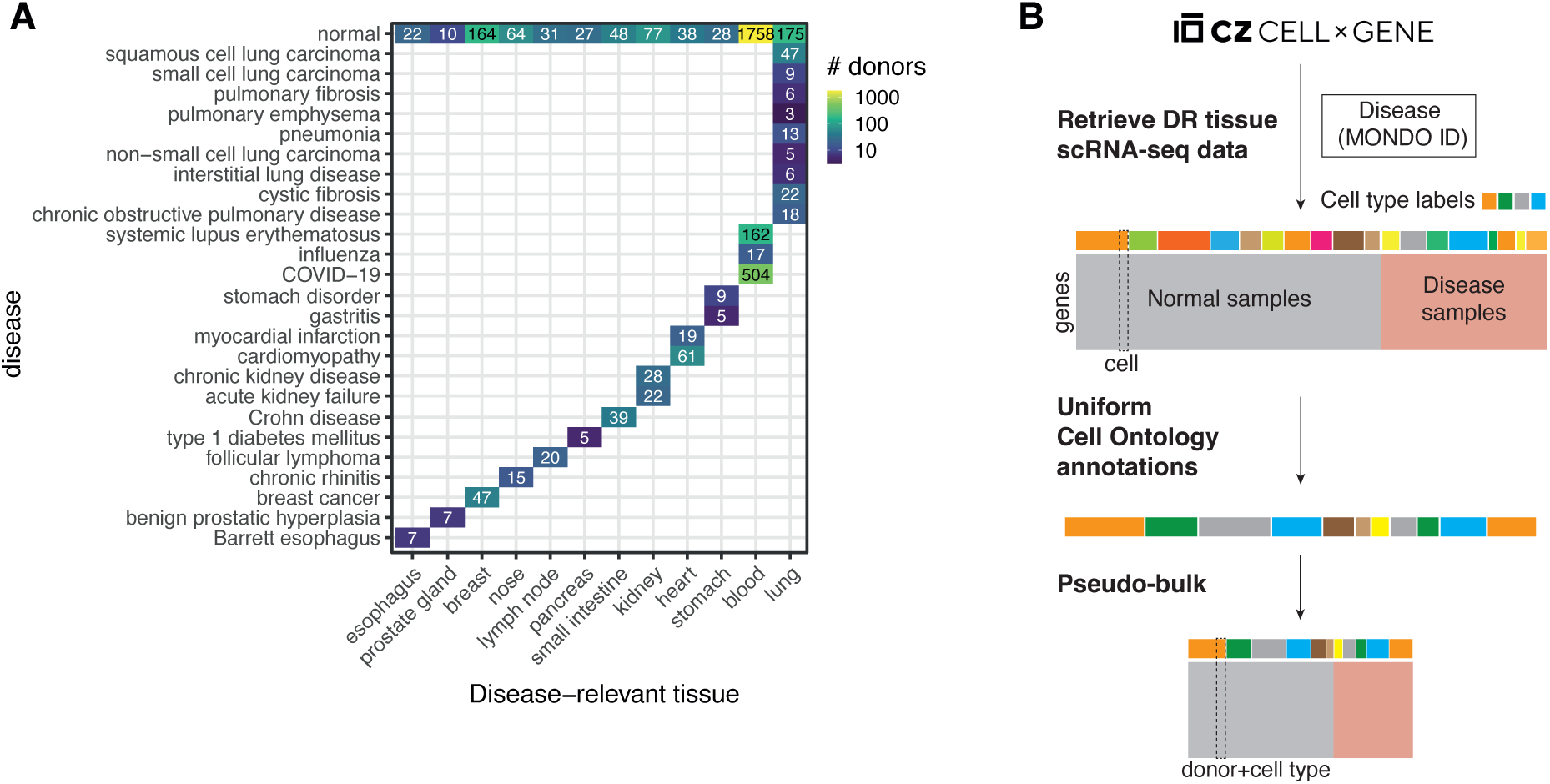
Single-cell RNA-seq dataset selection and pre-processing. (A) Overview of diseases and tissues in scRNA-seq dataset. Table of disease-relevant tissue of samples (x-axis) and disease condition (y-axis) for all scRNA-seq data considered in this study. The number and color of each square indicates the number of individuals for whom scRNA-seq data are available. The availability of data from healthy individuals is shown in the top row (disease condition: normal). (B) Illustration of selection and pre-processing steps for scRNA-seq datasets from CZ CellxGene Discover database (DR: disease-relevant).

For each disease, we identified cell type specific and disease cell specific genes with highly variable gene (HVG) selection and differential expression (DE) analysis, aggregating mRNA counts across cell types and donors (*Figure 1B*, see Methods). With this analysis across 25 diseases, we annotated 27780 gene-disease (G-D) pairs as cell type specific and 49253 G-D pairs as disease cell specific (Supplementary Figure 2). To explore associations between scRNA-seq support and clinical success, we extracted information about targets of drugs approved or in trial from the filtered Citeline Pharmaprojects table used by Minikel et al. [18]. This contrasts with a previous version of our analysis, where the Open Targets dataset was used as the main source of information on clinical targets (see Discussion and Methods for more details on the differences between clinical databases). The current dataset includes evidence for 2663 G-D pairs from drug development programs, of which 1648, 1208, 336 and 145 pairs have respectively reached phase I, phase II, phase III and market launch stages. We then computed the odds of reaching each phase of clinical development, with or without support from scRNA-seq data (see Methods, Supplementary Table 3). Of note, this analysis is disease-specific: we count successful G–D pairs with corresponding scRNA-seq support from analysis of healthy and diseased individuals in the disease-relevant tissue. For example, a gene that is cell type specific in oesophagus is not considered as scRNA-seq supported for pulmonary fibrosis.

In order to limit the analysis to only those genes expressed at a sufficient level in the disease relevant tissue, the association testing was restricted to highly variable genes (see Methods). Amongst all genes tested, we observed that scRNA-seq support both from cell type specificity and disease cell specificity significantly increase the odds of the cognate drugs to reach clinical phase I to III (Figure 3A-B). Since not all highly variable genes expressed in the disease relevant tissue are druggable, this result might indicate that cell type and disease cell specific targets are simply more likely to enter clinical development. Therefore, we next asked whether these features influence the overall likelihood of progressing to phase II and III for targets already in preclinical stages of drug development. To this aim, we restricted our analysis to the G-D pairs under active investigation by pharmaceutical companies, either at preclinical or clinical stages, as annotated in the Pharmaprojects data. Our results show that cell type specific targets are significantly more likely to reach phase I and II compared to non-cell type specific targets. However, the association is not significant for targets to reach phase III (Figure 3C). Conversely, disease cell specificity does not significantly increase the odds of reaching phase I, II or III (Figure 3D). Surprisingly, we also observed weak negative association between both cell type and disease cell specificity, and drug launch. As a positive control, we compared the odds observed with scRNA-seq support in this subset of G-D pairs with those derived from genetic support of the target, as defined by Open Targets direct genetic association score [1] and by Minikel et al [18]. As previously observed [16–18], when testing amongst highly variable ones, genes with genetic support are associated with clinical success, and the association is stronger for G-D pairs reaching phase III and launch. However, the association between genetic support and clinical success is no longer significant when the analysis is restricted to targets in preclinical or clinical stage. However, the odds of launch are markedly higher than those observed with scRNA-seq support. The discrepancy between these results and previous studies [16–18] can be explained by the smaller disease set used here, which is also biased towards immune diseases for which genetic evidence is known to be weakly associated with clinical progression [18].

**Figure 3.**
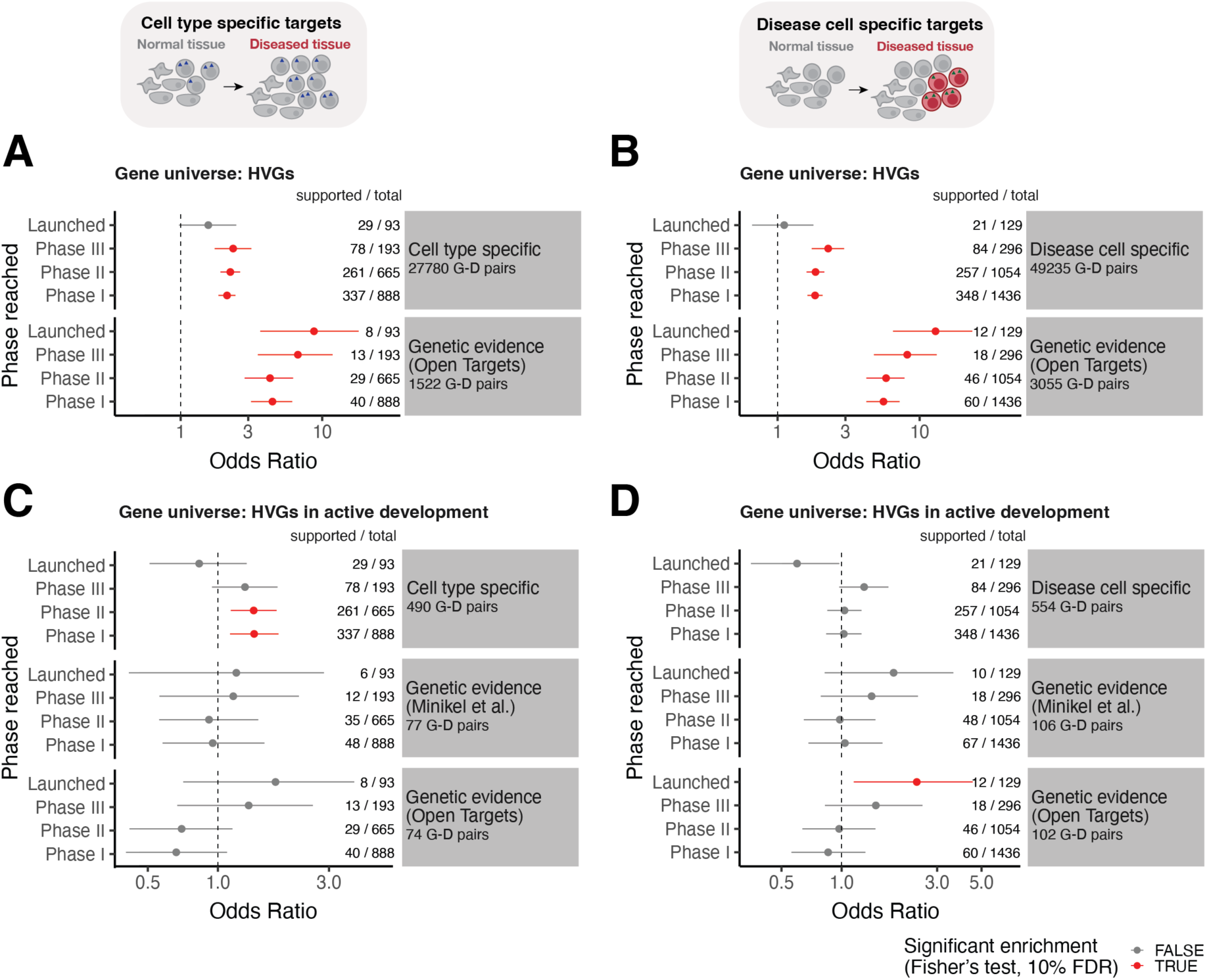
Association between omic-based evidence and target clinical success. Odds ratio for association between omic evidence (y-facets) and clinical phase reached (y-axis) for target-disease pairs for 25 diseases with available single cell data from CellxGene and disease ontology information based on Medical Subject Headings (MeSH) IDs. The left panels (A, C) show association testing for cell type specific targets. The right panels (B, D) show association testing for disease cell specific targets. For comparison between scRNA-seq support and genetic evidence, we show association with genetic evidence as defined by Minikel et al. (2024) and by Open Targets genetic association annotation. (A-B) Enrichment amongst all highly variable genes used for differential expression analysis; (C-D) Enrichment amongst highly variable genes that are under active investigation (using Pharmaprojects data processed by Minikel et al. 2024). For each test, the numbers to the right show the number of omic-supported targets over total targets that have reached each phase. The totals differ between test on cell type specific genes and disease cell specific genes because the set of HVGs used for the DE test is different (see Methods). The error bars denote 95% confidence intervals of the odds ratio. Points in red indicate cases where the enrichment for successful targets was statistically significant (Fisher’s exact test, Benjamini-Hochberg adjusted FDR < 10%). The dotted line denotes Odds Ratio = 1 (no enrichment). Genetic evidence as defined by Minikel et al. is shown only in C-D panels since annotation of genetic evidence in this database was available only for the gene-disease pairs in preclinical or clinical development.

Results are comparable also in progression analysis considering only targets that have entered a specific clinical phase (Supplementary Figure 3). Also in this case, we observe a weak but significant association of cell type specificity with progression through phase I (OR = 1.39, adj. p-val = 0.0078), while genetic support does not show statistically significant association with progression through the later phases of clinical development. Stratifying by disease, we confirmed that these trends are not driven by a specific disease or disease-relevant tissue (Supplementary Figure 4).

Taken together, these results suggest that cell type and disease cell specificity are features of targets that enter clinical development but are not predictive of success in late phases of clinical development. However, cell type specificity is robustly associated with early-stage success, suggesting that a cell-targeted mechanism may be associated with lower toxicity.

We next proceeded to the validation of these results on a larger set of indications. To expand the analysis to a larger number of diseases we focused on the 227 diseases in the Pharmaprojects data for which a single disease relevant tissue could be identified amongst the eleven available in CZ CellxGene database (Figure 4, Supplementary Table 4). Of note, in this analysis the cell type specificity was derived only from healthy donors and not from patients’ data.

**Figure 4.**
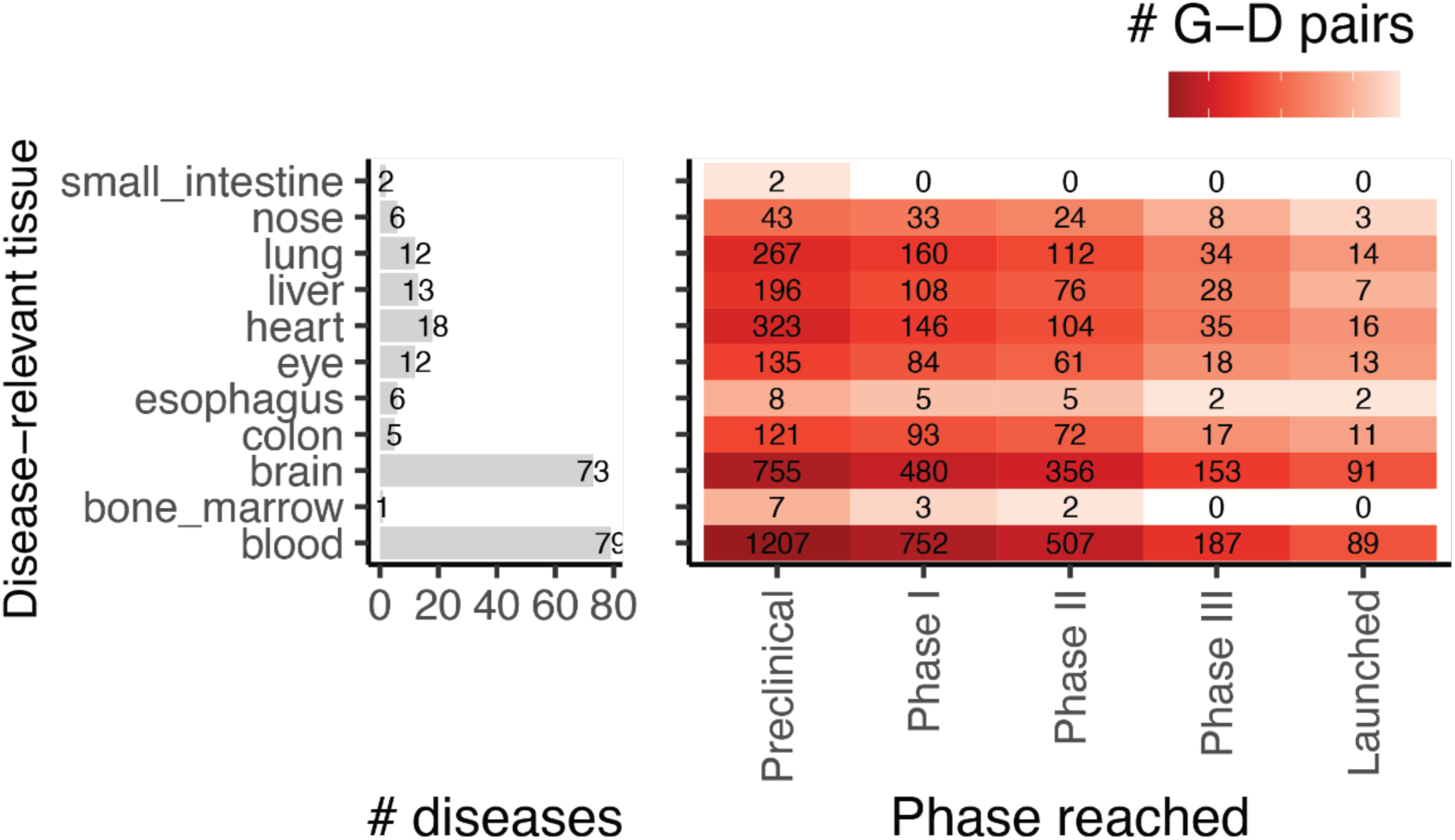
Overview of tissues and diseases considered for expanded cell type specificity analysis. The barplot shows the number of diseases considered for each Disease-relevant (DR) tissue from filtered Pharmaprojects table. The heatmap shows the total number of G-D pairs reaching each clinical phase for each DR tissue. Here the” Preclinical” category denotes the size of the gene universe for each DR tissue.

In this set of G-D pairs, the fraction of cell type specific targets in phase I and II of clinical development is higher than targets supported by genetic evidence. Conversely, the fraction of targets of launched drugs with genetic support is substantially higher than those supported by cell type specificity (Figure 5A). The odds ratio analysis replicates what we observed in the smaller disease set, whereby cell type specificity increases the odds of access to early stages of clinical development, but not of approval (Figure 5B, Supplementary Table 5). In this larger set of diseases we could also replicate the association between genetic evidence and progression previously reported by Minikel et al [18].

**Figure 5.**
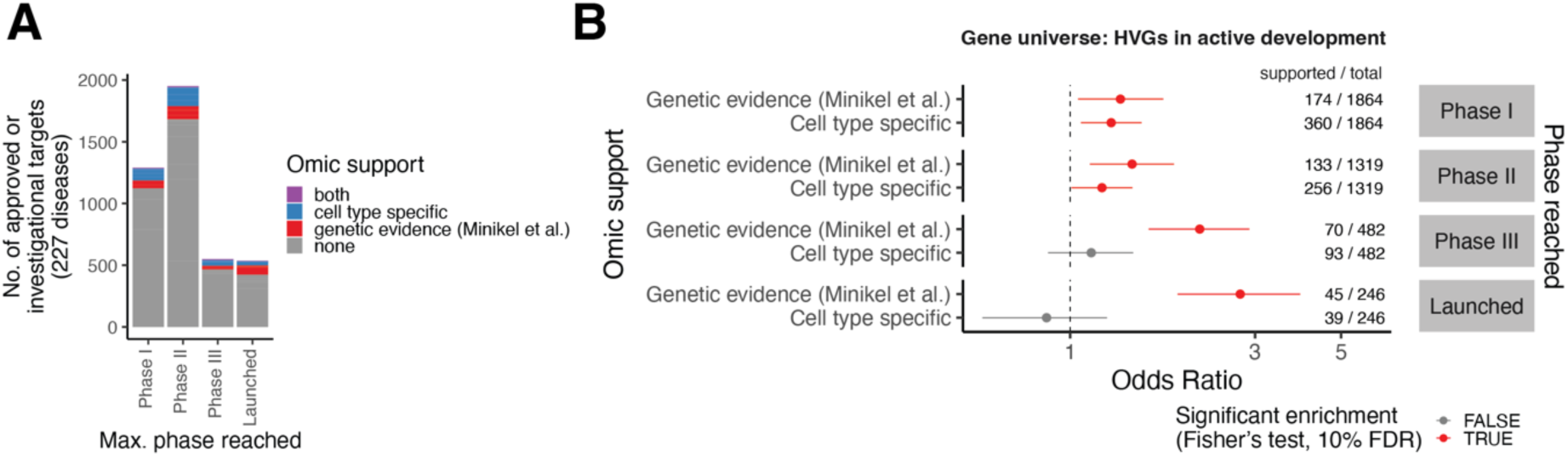
Expanded cell type specificity analysis on 227 diseases in filtered Pharmaprojects table and disease-relevant tissue data in CellxGene database. (A) Barplot of total number of G-D pairs (y-axis) by maximum clinical phase reached. G-D pairs supported by cell type specificity, genetic evidence or both are colored. (B) Odds ratio for association between omic evidence (y-axis) and clinical phase reached (y-facets) for target-disease pairs for 227 diseases. For comparison between scRNA-seq support and genetic evidence, we show association with genetic evidence as defined by Minikel et al. (2024). For each test, the numbers to the right show the number of omic-supported targets over total targets that have reached each phase. The error bars denote 95% confidence intervals of the odds ratio. Points in red indicate cases where the enrichment for successful targets was statistically significant (Fisher’s exact test, Benjamini-Hochberg adjusted FDR < 10%). The dotted line denotes Odds Ratio = 1 (no enrichment). Genetic evidence as defined by Minikel et al. is shown since annotation of genetic evidence was available only for the gene-disease pairs in preclinical or clinical development.

When we stratified the analysis by tissue, the association between cell type specificity and progression in early stages results to be general and not driven by any specific disease area (Supplementary Figure 5). Interestingly, when we focused on progression from phase I to II, we could identify specific disease areas, such as immune diseases where blood was annotated as disease-relevant tissue, where cell type specificity is a stronger indicator than genetic support, despite the fact that more genes have genetic support than scRNA-seq support (59 G-D pairs with genetic support and 37 G-D pairs with cell type specificity support, of 936 total G-D pairs passing phase I). Additionally, cell type specificity showed a weak, albeit not significant, negative association with progression from stage I to II for lung and colon diseases. We speculate that this might be due to our empirical definition of tissues deemed to be disease-relevant. Respiratory disorders might indeed affect specific or multiple sections of the airways, and a more refined definition of tissues involved might lead to a more effective definition of cell type specificity. Similarly, the colon was used in the analyses as a disease-relevant tissue for gastrointestinal disorders even though Crohn’s disease lesions can occur anywhere in the gastrointestinal tract, from the mouth to the anus. Moreover, both Crohn’s disease and ulcerative colitis can have several extra-intestinal manifestations in the eyes, hepatobiliary system, joints, and other tissues.

## Discussion

The ever-expanding use of scRNA-seq data to study disease, is accompanied by an increasing interest in quantifying the extent to which these data can support the drug discovery process, by identifying genes with increased probability of success as therapeutic targets early on. Our analysis is aimed at assessing whether cell type-specific features commonly derived from scRNA-seq studies are associated with increased success of targets to progress through preclinical and clinical development. We tested two distinct patterns of cell type specific expression: cell type specific expression in disease-relevant tissue (cell type specificity) and cell type specific over-expression in disease-relevant tissue from disease patients compared to controls (disease cell specificity).

Our results show that cell type specificity is associated with increased likelihood of entering clinical development and passing phase I, making it valuable information for target prioritization in the drug discovery process. Importantly, cell type specificity doesn’t require patient data, only harmonized cell type labels, and cell type-specific gene annotations for each tissue could be systematically computed and shared through open resources like the Human Cell Atlas Data Portal (data.humancellatlas.org) or the CZ CellxGene database [22]. Our analysis also points to the importance of the accurate definition of tissues defined as relevant for each given disease, something that was proved challenging, especially for complex diseases. We expect that complementary approaches such as heritability enrichment [24] could be used to improve this analytical framework by more accurately and systematically matching diseases and their cognate involved tissues.

Our analysis found no evidence linking cell type or disease cell specificity of a target with the likelihood of drug approval. Although we followed established best practices and utilized a large dataset, we believe these results should not be considered as conclusive with regards to the role of single-cell expression data in drug development. Several considerations can be made on the limitations of the results presented, and on potential areas for future analysis, suggesting that further research is necessary to fully understand this relationship.

First, clinical evidence data for target-disease pairs is sparse, with notable inconsistencies between data sources. For example, comparing the filtered Pharmaprojects dataset [18] with annotations of target-disease pairs from Open Targets [1], there are notable discrepancies in number of target-disease pairs associated to each clinical phase for the same set of diseases, which leads to different results for omic evidence association analysis (Supplementary Figure 6A-B). Not only is there a difference in size between the curated Pharmaprojects and Open Targets datasets, but also many G-D pairs are annotated in a specific clinical phase by one source only (Supplementary Figure 6C). The differences are especially marked amongst what are considered targets of approved drugs.

Also, publicly available data on clinical trials are often sparse and full information is not available regarding the reasons why programs failed. Of the 74 G-D pairs indicated as failing in phase III by Minikel et al. [18], only 13 pairs had annotations regarding the reasons for their cessation from a recent study on this topic [25]. Considering G-D pairs from Razuvayevskaya et al. [25], the majority of G-D pairs not passing phase III with single-cell support fail because of general “strategic” reasons (Supplementary Figure 7). Larger sets of gene-disease pairs and more granular information would be needed to more accurately determine the association between cell type or disease cell specificity with lack of demonstrated efficacy.

Furthermore, this work relies heavily on harmonized collections of scRNA-seq data, such as the CZ CellxGene Discover database. As a result, in certain diseases our analysis was constrained by the availability of tissue samples from only a small number of individuals (Figure 2A), and we found that with a larger patient cohort we could detect more disease cell specific genes (Supplementary Figure 10). Another potential limitation lies in the use of Cell Ontology-based cell type labels [23], which rely on annotations provided by curators upon submission to the database. These might penalize the accuracy of cell annotations, with the same label applied to transcriptionally distinct cells or different labels for identical cell types. While label harmonization partially addresses some issues, it leads to coarser annotations and penalizes the accurate labelling of rare, tissue-specific subpopulations.

Importantly, while we found that cell type specificity is an informative feature for success in early stages of clinical development, more sophisticated analyses on scRNA-seq data might be needed to identify a cell-centric diagnostic that is predictive of drug efficacy. We define disease cell specificity using a naïve cell type-level differential expression analysis, where technical effects are only partially mitigated. This could lead to false discoveries that could be mitigated by refined experimental design and using more robust statistical methods to recover expression differences in scRNA-seq data [26]. Additional improvements might come from the pre-selection of disease-relevant cell types to derive cell type specificity. This could be achieved through differential cell abundance analyses [27,28], other supervised methods for multi-condition scRNA-seq data [29–31] or heritability enrichment [32,33]. Furthermore, methods to infer differentiation trajectories [34,35], cell-cell interactions [36,37], regulatory networks [38], and immune repertoires [39] from single-cell genomics provide additional unexplored space for novel targets. Our work provides a framework and baseline to assess the potential impact of alternative single-cell data analysis methods and modalities on target discovery.

Finally, it is worth considering that retrospective analyses such as the one presented here focus on historical data. This implies that therapeutic mechanisms in diseases where cell type-specific analyses have the greatest impact may not have been fully explored or exploited. For example, we found that a significant proportion of cell type-specific and disease-specific genes are considered tractable by therapeutic agents (Supplementary Figure 8). Notably, cell type-specific genes are particularly enriched in membrane-bound proteins, which are promising targets for certain therapeutic modalities such as antibody-based therapies. In addition, cell-specific therapies might have the greatest benefits in patient subsets identifiable by cell-driven precision medicine approaches, which are rarely implemented today.

In summary, our work demonstrates that currently standard single-cell analyses can aid in prioritizing druggable and safe targets. However, more refined data curation and analytical methods are necessary to support the later phases of clinical development with evidence-based approaches. We hope this serves as a foundation for future research focused on leveraging emerging tissue profiling technologies for target discovery.

## Methods

### Single-cell RNA-seq data collection from CZ CellxGene Discover platform

To select a set of diseases and scRNA-seq datasets, we downloaded cell- and dataset-level metadata for all *H.Sapiens* datasets from the CZ CellxGene Discover database, using the *cellxgene_census* python API (census version: 2023-07-25) [22]. Disease-relevant (DR) tissues were manually annotated for the 58 disease terms in the database. We excluded datasets profiled with targeted scRNA-seq assays (BD Rhapsody), inDrop and STRT-seq. We further excluded fetal samples, based on Human Developmental Stage Ontology [40], where available, and by manual curation for 12 datasets where stages were annotated as “unknown”. 10 disease terms were grouped into 4 broader terms (Supplementary Table 1).

After curation, 25 disease terms were retained for association analysis. Reasons to exclude diseases included: missing overlapping disease terms in clinical databases (filtered Citeline Pharmaprojects or Open Targets), missing data from DR tissue, data available from less than 3 donors with the disease, download errors (see Supplementary Table 1 for a complete list of diseases and reasons to exclude from analysis). After selecting suitable datasets, for each disease we downloaded full transcriptome gene expression profiles for all cells from the DR tissue from healthy donors and disease patients, as well as cell type labels (Cell Ontology terms [23]) and sample-level technical metadata (scRNA-seq assay and suspension type, Supplementary Figure 11).

To ensure consistency in granularity of cell type annotations across studies, we implemented a rollup procedure on the Cell Ontology tree, by relabelling cells with parent terms if a given term is a descendant of another term in the dataset (see example outcome in Supplementary Figure 1). For each term, the search for parent terms was limited only to a level of depth in the ontology tree given by the total number of ancestors of the term divided by a factor of 5. For example, if a term had 20 ancestors in the ontology tree, we searched for the 4 closest parent terms in the dataset for relabelling. We recognize that this step reduces the resolution of cell type annotations, yielding broader and partially redundant annotation labels. However, it mitigates the need for batch correction, clustering, and manual cell type annotation across 30 datasets. We defined the cell type labels used after roll-up as *high-level cell type annotations*.

For the analysis of cell type specificity in healthy DR tissue on an expanded set of diseases (Figure 4), we considered all tissues in the CZ CellxGene database for which data from at least 3 healthy individuals was available.

### Differential expression analysis and extraction of scRNA-seq supported gene-disease pairs

We identified cell type specific and disease cell specific genes for each disease using differential expression (DE) analysis.

For each disease dataset, we aggregated cell-level gene expression profiles summing counts and size factors (total counts per cell) by donor and high-level cell type annotations (hereafter, pseudo-bulks), following best practice recommendations for DE analysis on scRNA-seq data [26,41]. Only cell types found in at least 3 healthy donors (and 3 disease donors for disease cell specificity analysis) were included in DE testing. To identify cell type specific genes, we selected pseudo-bulks from healthy donors from the disease-relevant tissue and we tested for DE between pseudo-bulks of one cell type against all other cell types. To identify disease cell specific genes, for each cell type we tested for DE between diseased donors and healthy donors. For each test, we selected the top 5,000 highly variable genes amongst considered pseudo-bulks, using the method implemented in the R package *scran* [42]. We tested for differential expression between groups with the *edgeR* quasi-likelihood test [43] using the implementation in the R package *glmGamPoi* [44]. In all tests, we modelled the number of cells per pseudo-bulk as a confounder, as well as suspension type (cell or nuclei) and scRNA-seq assay where possible (when the confounder was not perfectly collinear with the disease label). After DE analysis, we obtained the effect size (log-fold change, logFC) and Benjamini-Hochberg adjusted p-values for each tested gene in each tested cell type.

We annotated a gene-disease (G-D) pair as cell type specific when the gene is significantly over-expressed in at least one cell type compared to all other cell types in healthy disease-relevant tissue (adjusted p-value < 0.01, logFC > 5). The choice of logFC threshold was motivated by the observation that genes significantly over-expressed at lower log-fold changes are often ubiquitously highly expressed, while those at higher fold changes are genuinely cell type specific (*Supplementary Figure 9*). We annotated a G-D pair as disease cell specific when the gene is significantly over-expressed in disease in at least one cell type in disease-relevant tissue (adjusted p-value < 0.01, logFC > 0.5). The total number of supported G-D pairs for each disease is shown in Supplementary Figure 2.

### Drug development data

We used data on clinical progression for G-D from the Citeline Pharmaprojects database, as filtered, pre-processed and re-distributed by Minikel et al. [18]. Briefly, this includes monotherapy programmes added since 2000 annotated with a highest clinical phase reached and where a human gene target and a disease indication were available. The table was downloaded from the study supplementary materials. We selected this as our primary source of drug development data to ensure comparability with the latest retrospective analysis studies [18,45] and to use the annotation of targets considered for pre-clinical development, which is missing from alternative publicly-available resources. For the extended analysis, we assigned disease-relevant tissue for each indication, starting from the annotations of therapeutic areas by Minikel et al. Indications ‘oncology’, ‘congenital’, and ‘other’ (except for kidney and prostate disorders) were excluded from this extended analysis due to concerns about the ability to select an appropriate disease-relevant tissue. Digestive disorders were subdivided into those with liver, oesophagus, small intestine, or colon as their disease-relevant tissue. Hematologic disorders could be assigned blood, lymph node, or bone marrow. Respiratory disorders could be assigned nasal tissue or lung. Indications where a disease relevant tissue could not be identified were excluded, leaving 227 diseases for inclusion in the extended analysis (Supplementary Table 4). We decided to always assign a single tissue to each indication to avoid potential artifacts coming from analysing much larger set of gene-disease pairs for indications with multiple tissue assignments.

For comparison between sources of clinical evidence (Supplementary Figure 6), we collected data on clinical progression for G-D pairs from Open Targets, using two distinct sources:

-Open Targets (ChEMBL score): Open Targets direct association evidence was accessed via download from the Open Targets Platform (version 23.02) [1,46]. Downloads used for this analysis were the ‘Diseases’ and ‘Direct Associations by Type’ tables. Experimental Factor Ontology (EFO) disease terms used in Open Targets were mapped to their corresponding term in used in the CellxGene database (MONDO IDs) using the ontology tree available in the Open Biological and Biomedical Ontology Foundry (https://obofoundry.org/ontology/mondo.html). We annotated G-D pairs for which approved or clinical candidate drugs exist using the ChEMBL evidence score from the Open Targets Platform. Briefly, each G-D pair is assigned a score between 0 and 1 based on clinical precedence, then the score is down-weighted by half if the clinical trial has stopped early for negative results (no effect of the drug) or safety and side effects concerns. Following the ChEMBL evidence scoring in Open Targets (https://platform-docs.opentargets.org/evidence#chembl), we classified G-D pairs with a ChEMBL evidence score > 0.1 as safe (> phase I), pairs with score > 0.2 as effective (> phase II), and pairs with score > 0.7 as approved (> phase III). While we do not explicitly exclude gene-disease pairs supported by failed trials, the down-weighting in Open Targets ensured that targets failed in early clinical trials are excluded, and targets failed in phase III were at most classified as passing phase II.
-Open Targets (Razuvayevskaya et al. 2024): data processed by a recent study from Open Targets [25] was downloaded from HuggingFace (https://huggingface.co/datasets/opentargets/clinical_evidence).

Genetic association

We used two distinct annotations of G-D pairs with genetic support, both aggregating evidence for association of genes and rare and common variants from several sources:

-Direct genetic association score provided in Open Targets (https://platform-docs.opentargets.org/evidence) [1]. We classified as supported by genetics any G-D pair with genetic association score > 0.
-Annotation of genetically supported G-D pairs from Minikel et al. [18], including evidence from an expanded set of indications, based on similarities on Medical Subject

Headings (MeSH) ontology tree. These annotations are reported only for G-D pairs under active investigation by pharmaceutical companies present in the filtered Pharmaprojects data. Therefore, we did not use these annotations for enrichment analyses on the set of highly variable genes.

Both genetic annotations derive from aggregated analysis of multiple data sources including associations with both rare and common variants implicated by association studies, and have been used in different studies [25,45].

### Association between omic evidence and progression through clinical trials

To test for association between omic evidence (cell type specificity, disease cell specificity, genetic association) and clinical success (reaching clinical phase I, II, III or launch) we computed the odds ratio and Fisher exact test p-value under the null hypothesis that the true ratio between the odds of being a successful G-D pair with omic support and of being successful without support is 1. In all association tests, drug indications for clinical success and data for omic support are aligned by disease. To compute odds ratios, 95% confidence intervals and p-values, we used the odds ratio calculation implementation in the python package *scipy* [47]. In all figures and tables, we report FDR corrected p-values using the Benjamini-Hochberg procedure. To enumerate the space of possible G-D pairs for odds ratios analysis, we used the following gene sets as “gene universes”:

-HVGs: all highly variable genes considered for DE analysis for each disease. We test on this set as standard practice for enrichment analysis after DE testing, and to consider genes that are expressed at a sufficient level in the DR tissue, regardless of their druggability potential. We recognize this might lead to a conservative estimate of association for cell type specificity, where highly variable gene selection between cell types should already be selecting weakly cell type specific genes. However, we observe that several HVGs selected for cell type specificity analysis are frequently ubiquitously expressed and cell type specific only at very low fold changes (*Supplementary Figure 9*).
-HVGs under active investigation: the intersection between genes considered for DE analysis and genes that are under active investigation by pharmaceutical companies for each disease, as annotated in the filtered Pharmaprojects dataset (see “Drug development data”).

## Data availability

All scRNA-seq data analysed in this study is available via the CZ CellxGene Discover database and CxG Census API (https://chanzuckerberg.github.io/cellxgene-census/, version: 2023-07-25). Data on clinical precedence for known drugs for each target-disease pair, as well as gene-disease genetic association scores, was obtained from the supplementary materials of Minikel et al. 2024 [18] and from Open Targets (version 23.02, https://platform.opentargets.org/downloads/data). Processed datasets and analysis outputs are available as supplementary tables and via figshare (doi:10.6084/m9.figshare.25360129) and through our code repository (https://github.com/emdann/sc_target_evidence).

## Code availability

All code to reproduce data downloads, processing and analysis is available at https://github.com/emdann/sc_target_evidence.

## Acknowledgements

We thank Jeffrey Greve and members of the Teichmann group for valuable discussions on this project. We thank Matt Nelson, Robert Scott, Daniel Seaton, Nicola Richmond, Rogier Hintzen, Dane Corneil and David Hulcoop for sharing critical insights and feedback on the first version of this analysis. ED, KBM and SAT acknowledge Wellcome Sanger core funding (WT206194).

## Author contributions

ED, ET, RE, GG, VS, EdR and SAT conceptualized the study. ET performed curation of Open Targets data and drug-level data analysis. ED performed curation and processing of scRNA-seq data, differential expression analysis, statistical analysis of association between omic evidence and clinical success, and disease-level target analysis. All authors interpreted the results. ED and ET made the figures. ED, ET, RE and EdR wrote the original manuscript draft. All authors edited and approved the final version of the manuscript. EdR and SAT supervised the work.

## Supplementary Figures

**Supplementary Figure 1:**
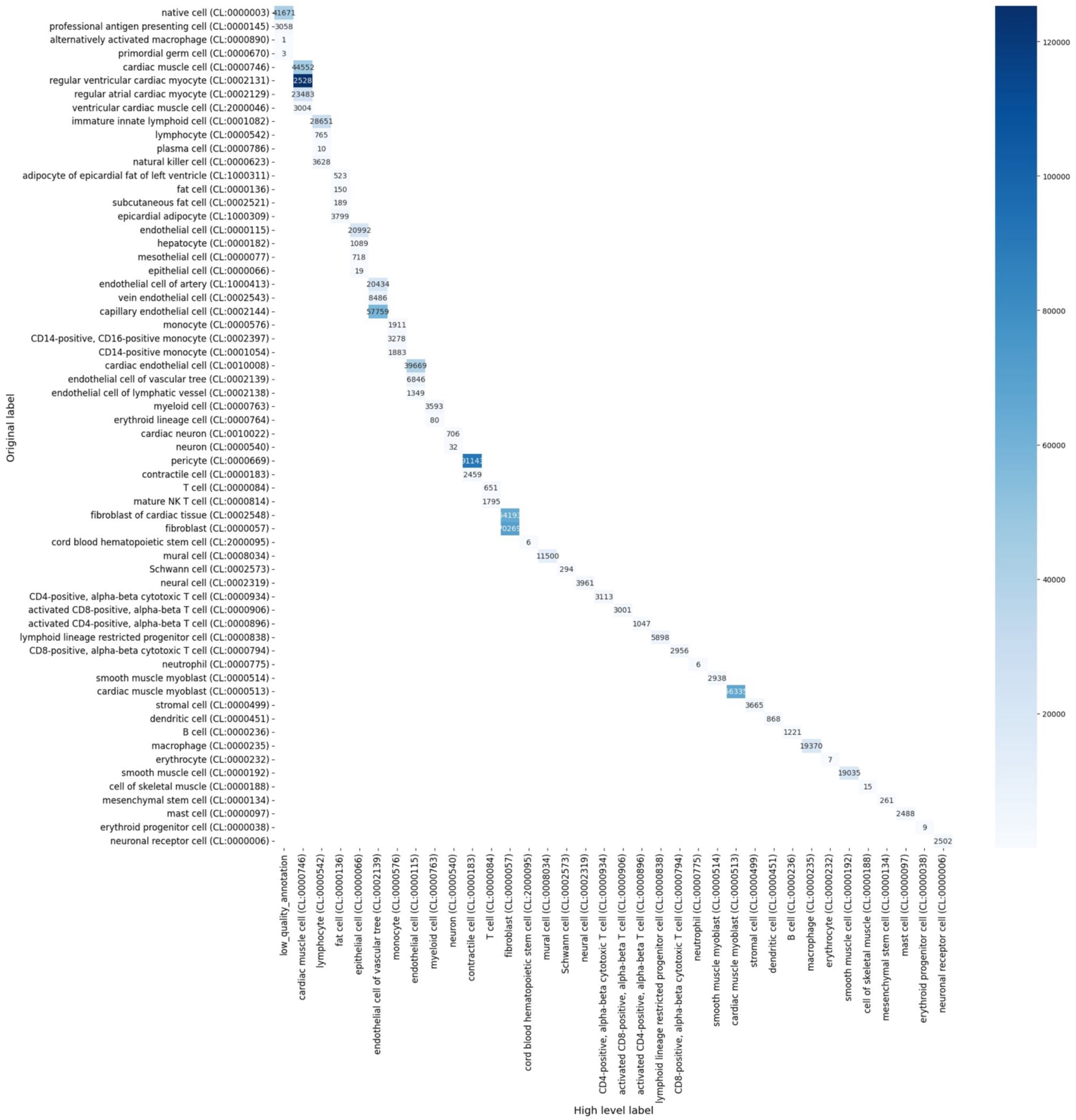
Example outcome of harmonisation of cell type annotations based on Cell Ontology. The y-axis shows the original Cell Ontology label used in CZ CellxGene database for the myocardial infarction dataset (disease-relevant tissue: heart) and the x-axis shows the updated label after label harmonisation. The heatmap color and number indicate the number of cells for each label.

**Supplementary Figure 2:**
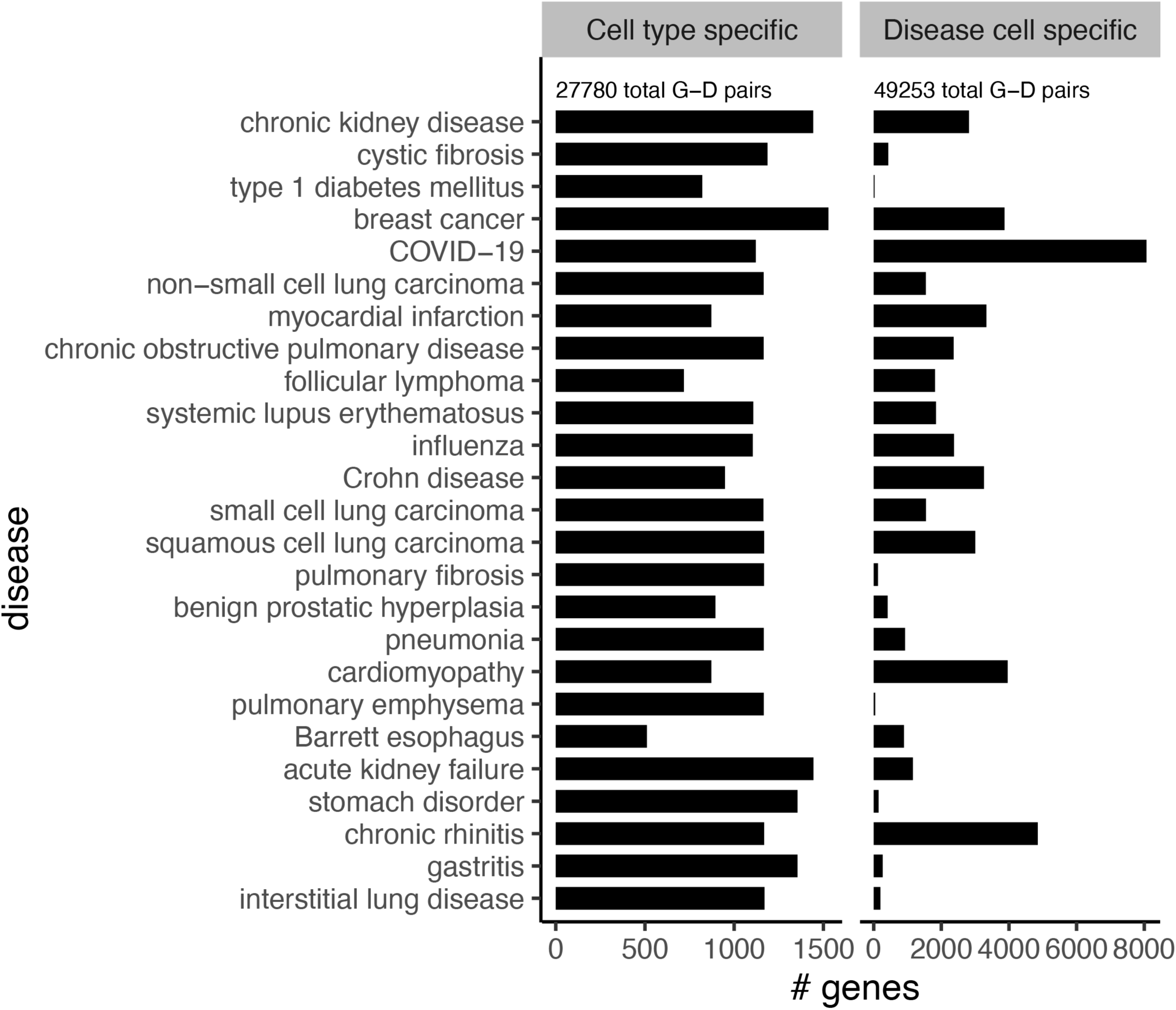
Number of clinically successful and supported targets per disease. Barplot of number of gene targets with scRNA-seq support by disease. The total number of G-D pairs for each class is reported above the bar plots.

**Supplementary Figure 3.**
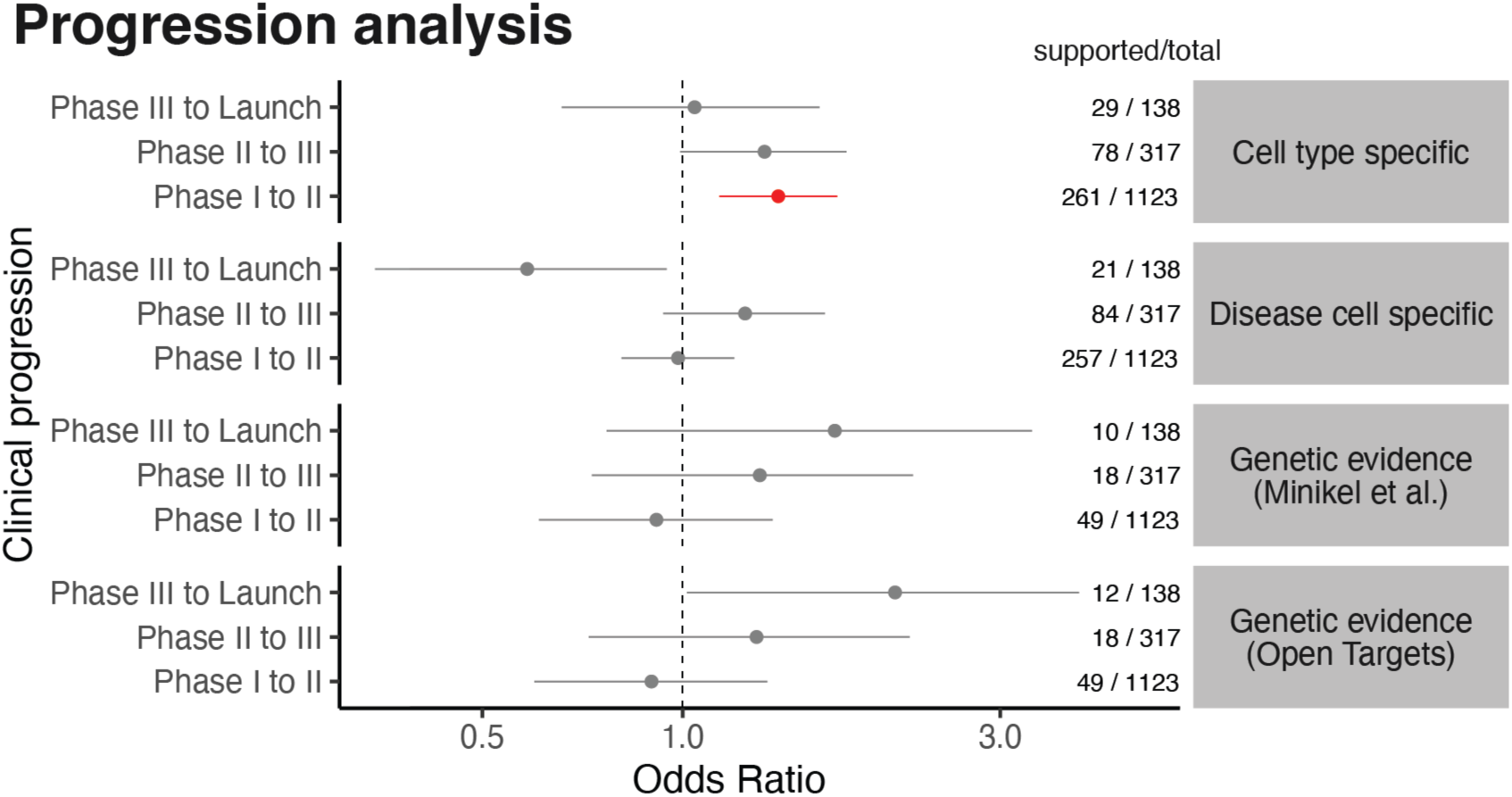
Progression analysis. Odds ratio for association between omic evidence (y-facets) and progression through clinical phases (y-axis) for gene-disease pairs for 25 diseases. For comparison between scRNA-seq support and genetic evidence, we show association with genetic evidence as defined by Minikel et al. (2024) and by Open Targets genetic association. For each test, the numbers to the right show the number of omic-supported targets over total targets that have reached each phase. The error bars denote 95% confidence intervals of the odds ratio. Points in red indicate cases where the enrichment for successful targets was statistically significant (Fisher’s exact test, Benjamini-Hockberg adjusted FDR < 10%). The dotted line denotes Odds Ratio = 1 (no enrichment).

**Supplementary Figure 4:**
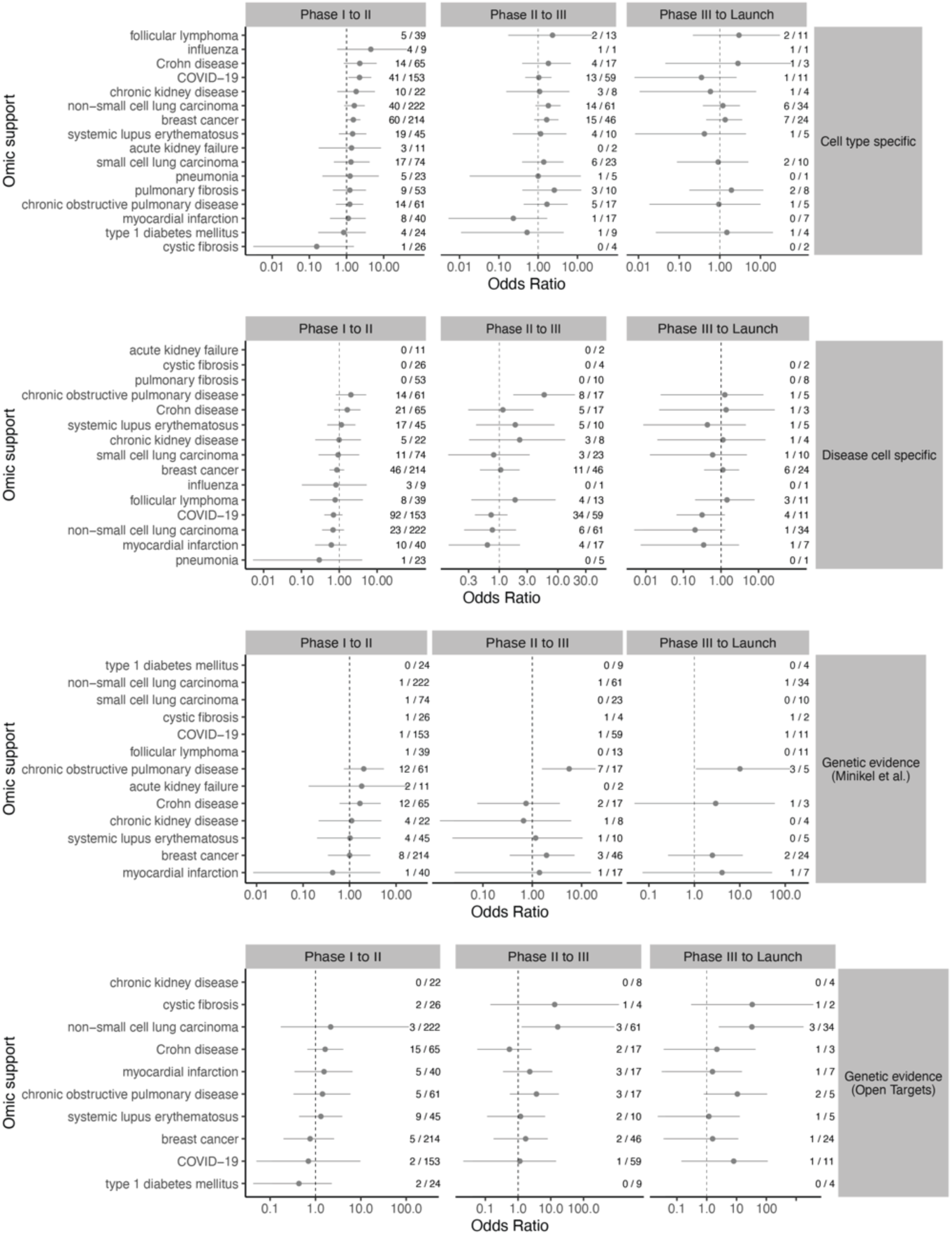
Association between omic support and clinical progression stratified by disease. Odds ratio (x-axis, in log10 scale) of association between clinical success of a target and scRNA-seq support (y-axis) computed stratifying by disease. Results are shown for diseases with at least 1 launched and 1 supported target. For each test, the numbers to the right show the number of omic supported targets over total successful targets. The error bars denote 95% confidence intervals of the odds ratio. Points in red indicate cases where the enrichment for successful targets was statistically significant (Fisher’s exact test p-value < 0.05). The dotted line denotes Odds Ratio = 1 (no enrichment).

**Supplementary Figure 5.**
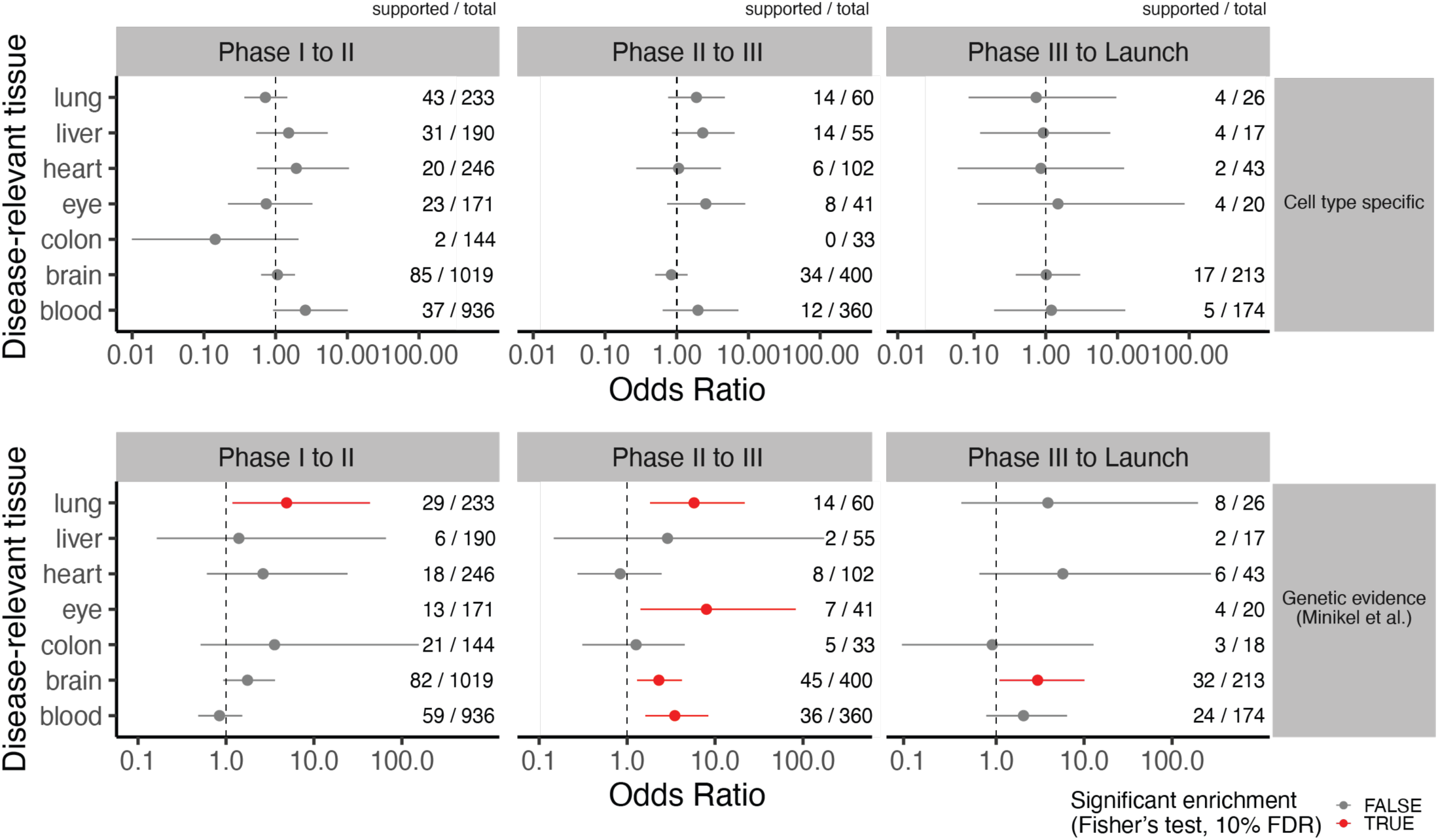
Association between cell type specificity and clinical progression stratified by disease-relevant tissue. Odds ratio (x-axis, in log10 scale) of association between clinical success of a target and scRNA-seq support (top panel) or genetic evidence (bottom panel. For each test, the numbers to the right show the number of omic supported targets over total successful targets. The error bars denote 95% confidence intervals of the odds ratio. Points in red indicate cases where the enrichment for successful targets was statistically significant (Fisher’s exact test p-value < 0.05). The dotted line denotes Odds Ratio = 1 (no enrichment).

**Supplementary Figure 6.**
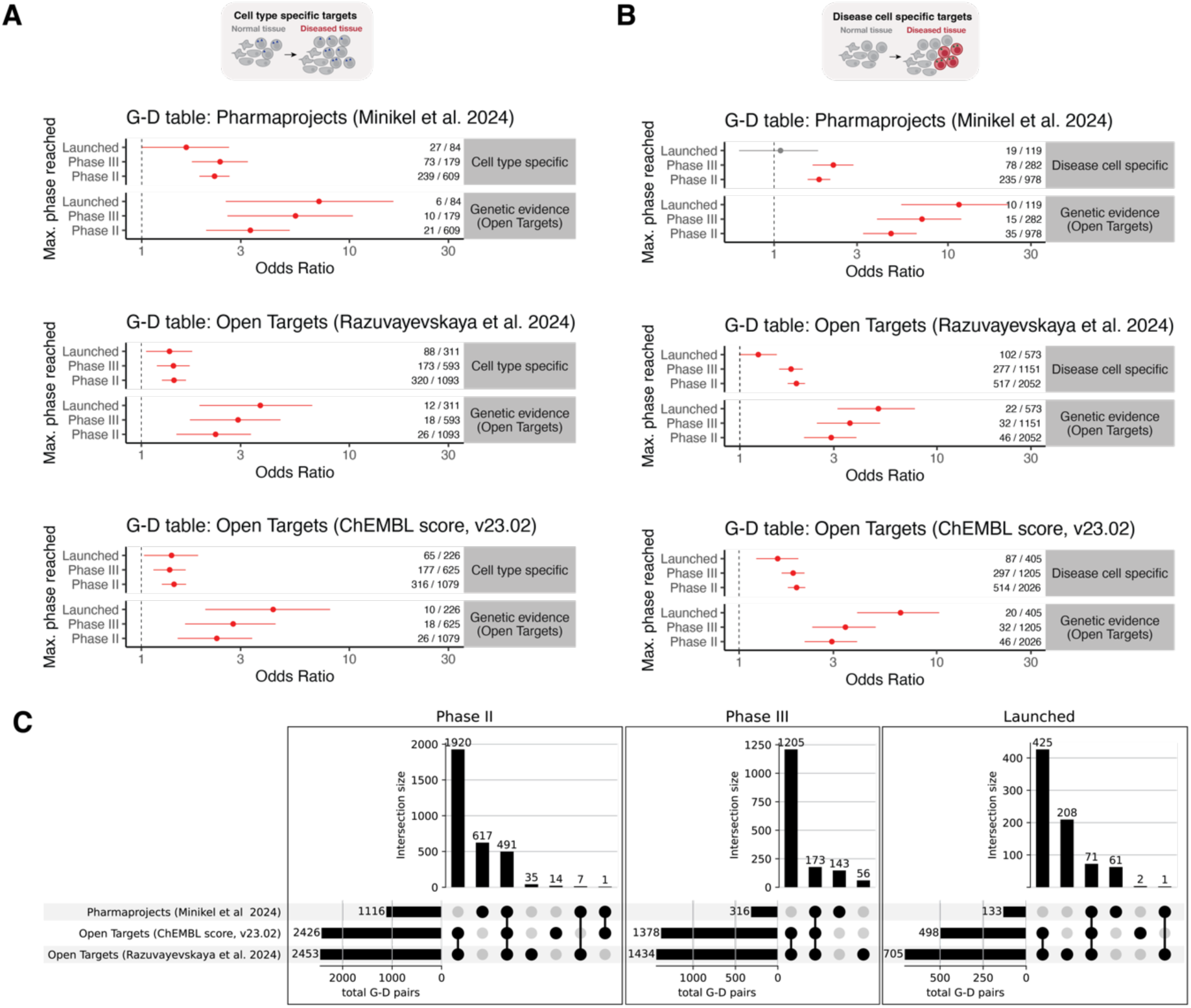
Comparison between sources of clinical evidence for gene-disease pairs. (A-B) Odds ratio for association between omic evidence (y-facets) and maximum clinical phase reached (y-axis), as in figure Figure 3A-B, computed using different datasets of clinical progression for G-D pairs: filtered Pharmaprojects data from Minikel et al. 2024, filtered Open Targets data from Razuvayesvskaya et al. 2024, unfiltered Open Targets data (v23.02) based on ChEMBL score. Results are shown for 24 diseases for which G-D pairs were available in all sources. This comparison is limited to tests within highly variable genes, since only the Pharmaprojects data annotates pairs of datasets in pre-clinical development. (C) Upset plots showing the intersection of G-D pairs that have reached each clinical phase between clinical evidence sources.

**Supplementary Figure 7:**
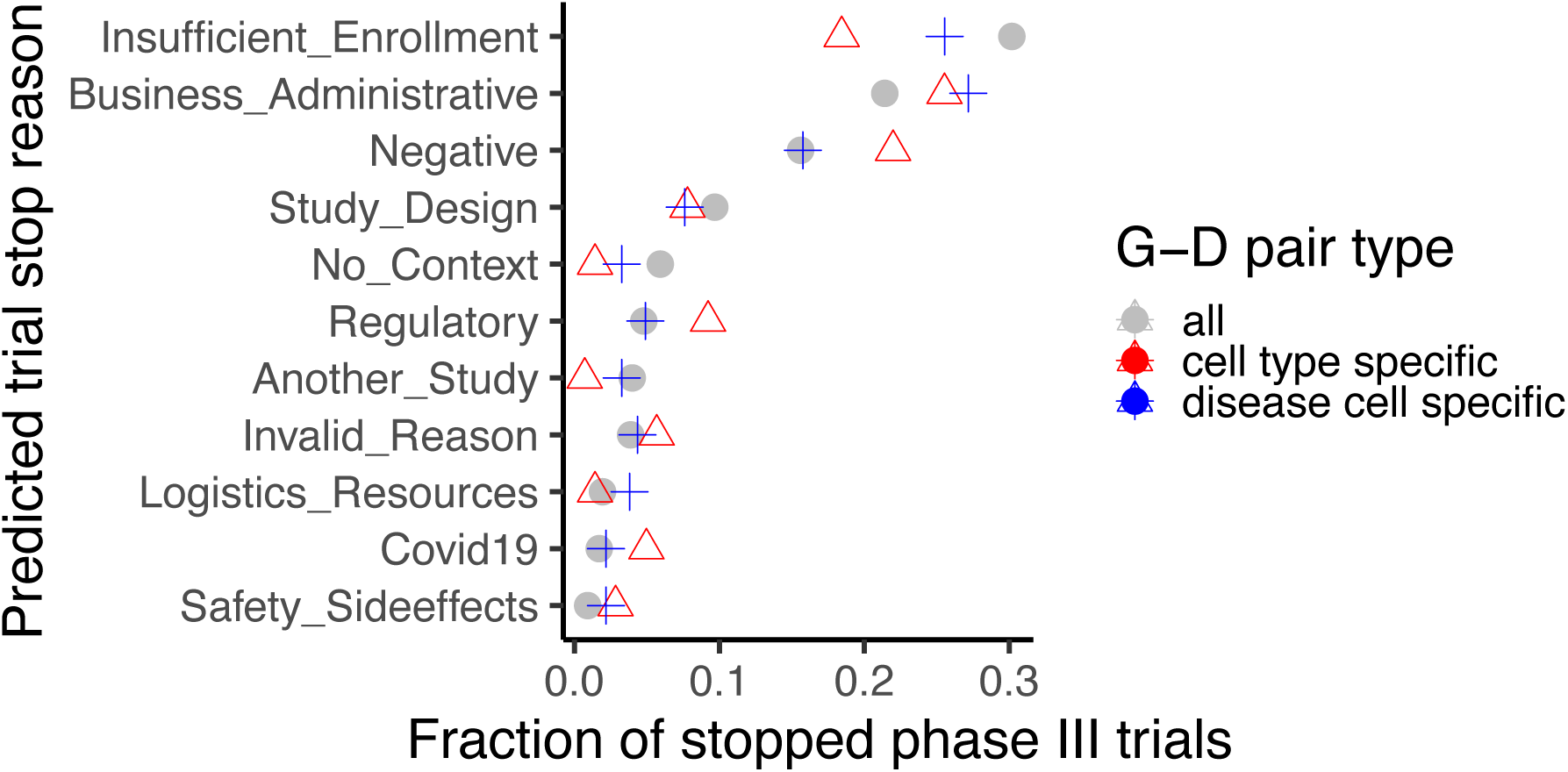
The proportion of trial failures due to various reasons, grouped by gene-disease (G-D) pair types. The x-axis represents the proportion of failures for each category, while the y-axis lists the different reasons for failure as predicted in a recent Open Targets study [25]. The distribution of failures highlights differences in failure reasons across G-D pair types, suggesting variability in trial success depending based on available target evidence.

**Supplementary Figure 8:**
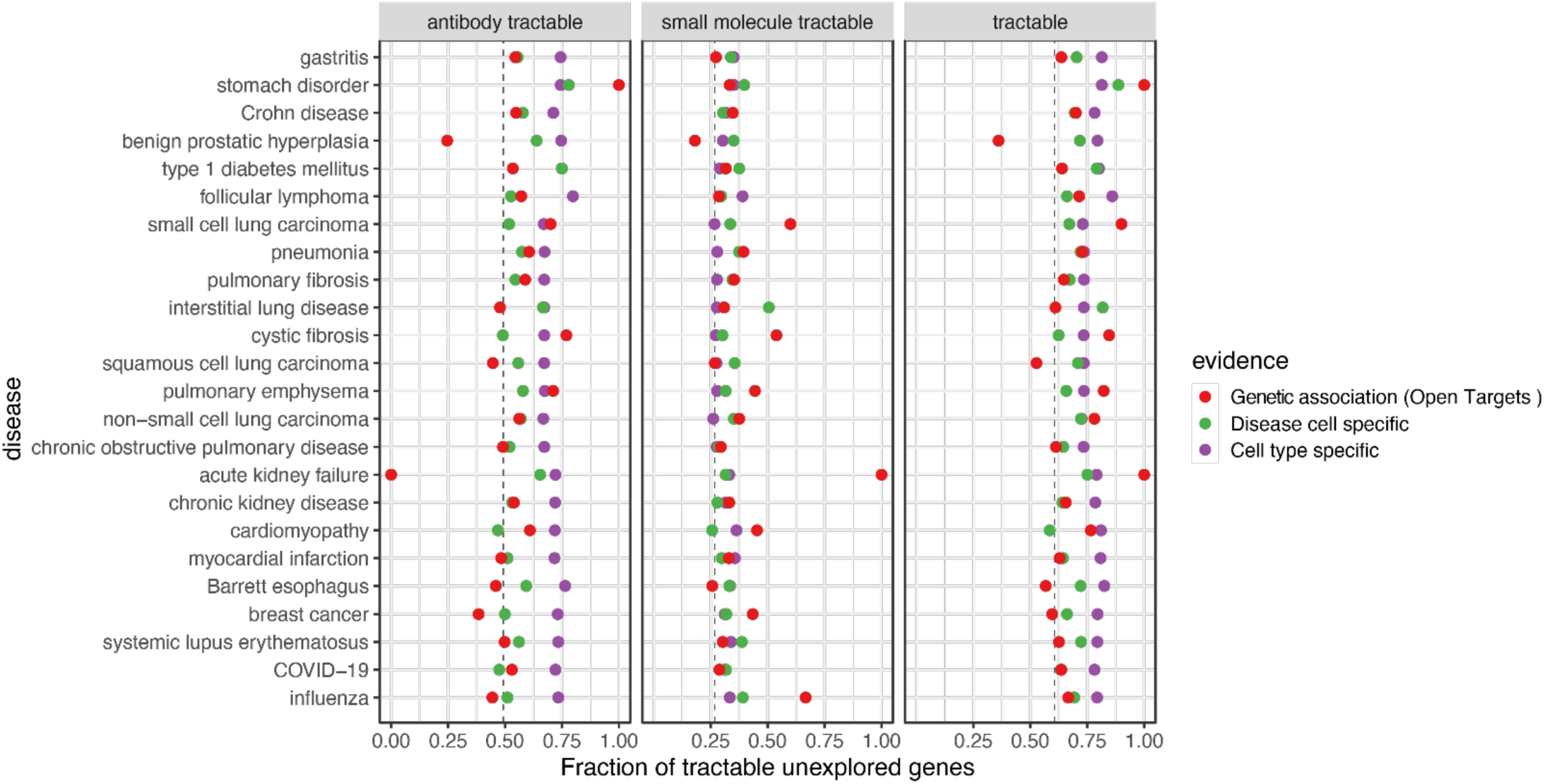
Tractability of unexplored targets across diseases. Scatter plot of fraction of tractable unexplored genes (x-axis) for 24 diseases (y-axis) for different classes of omic evidence (color). We consider three categories: antibody tractable, small molecule tractable, and tractable by either class of drugs. Dashed lines represent the fraction of tractable genes across all protein-coding genes. Diseases for which no gene with genetic evidence was found are not shown (n=3).

**Supplementary Figure 9:**
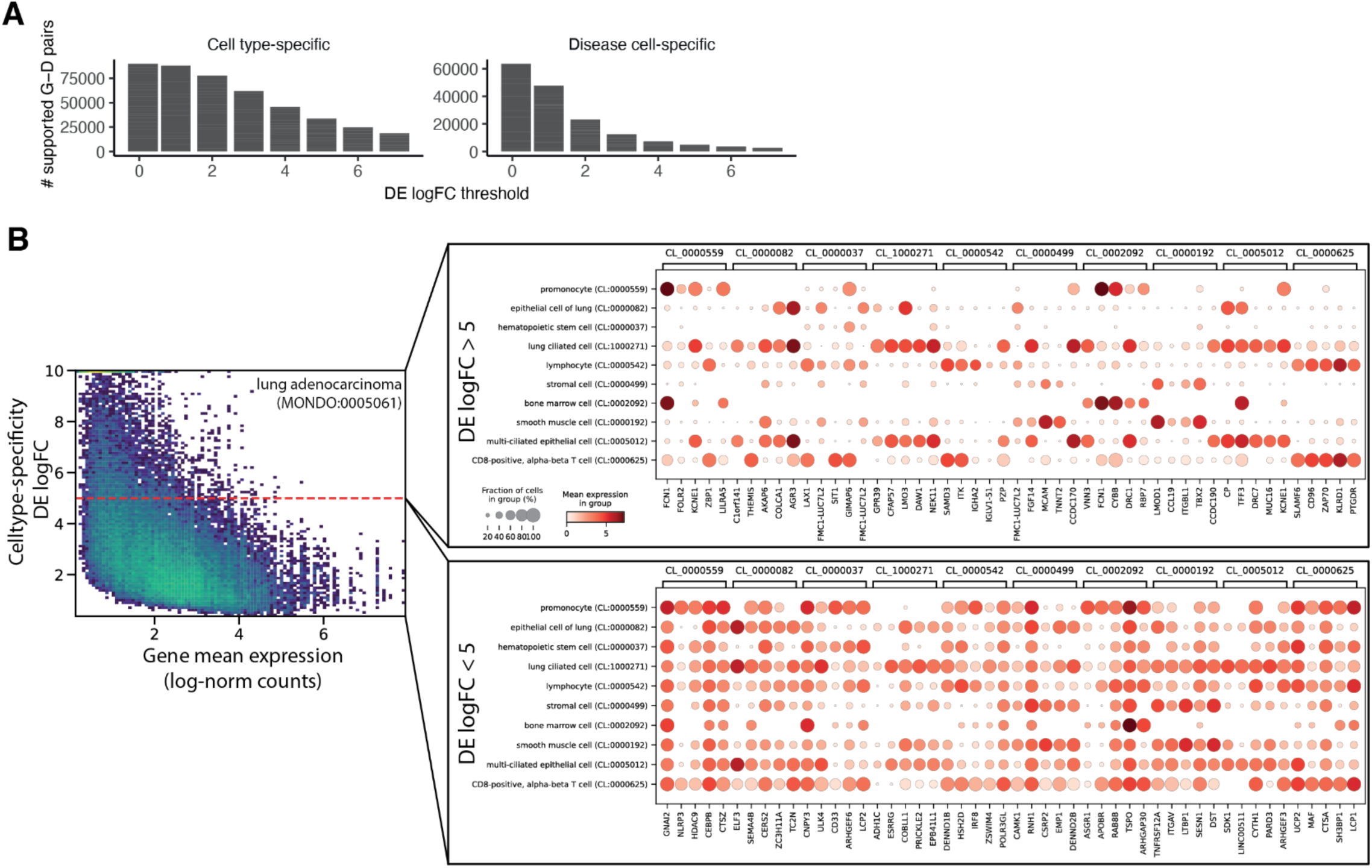
Analysis of parameters for definition of targets with scRNA-seq support. (A) Barplot of number of supported G-D pairs with increasing log-Fold Change (logFC) threshold on differential expression (DE) analysis results, for cell type specific genes (left) and disease cell specific genes (right). (B) Example from lung adenocarcinoma scRNA-seq data showing cell type specificity of candidate target genes at high DE log-fold changes. The left scatterplot shows the mean expression (log-normalized counts, x-axis) and DE log-fold change for one-vs-all test (y-axis) used for cell type specificity analysis for each significantly over-expressed gene (1% FDR). The dotplots to the right show the expression, in terms of mean (color) and cell fraction (size) for 5 randomly selected cell type specific genes detected in 10 lung cell types (the cell ontology term is indicated on top of the plots). The top plot shows significant genes with logFC > 5 and the bottom plot shows significant genes with logFC < 5.

**Supplementary Figure 10:**
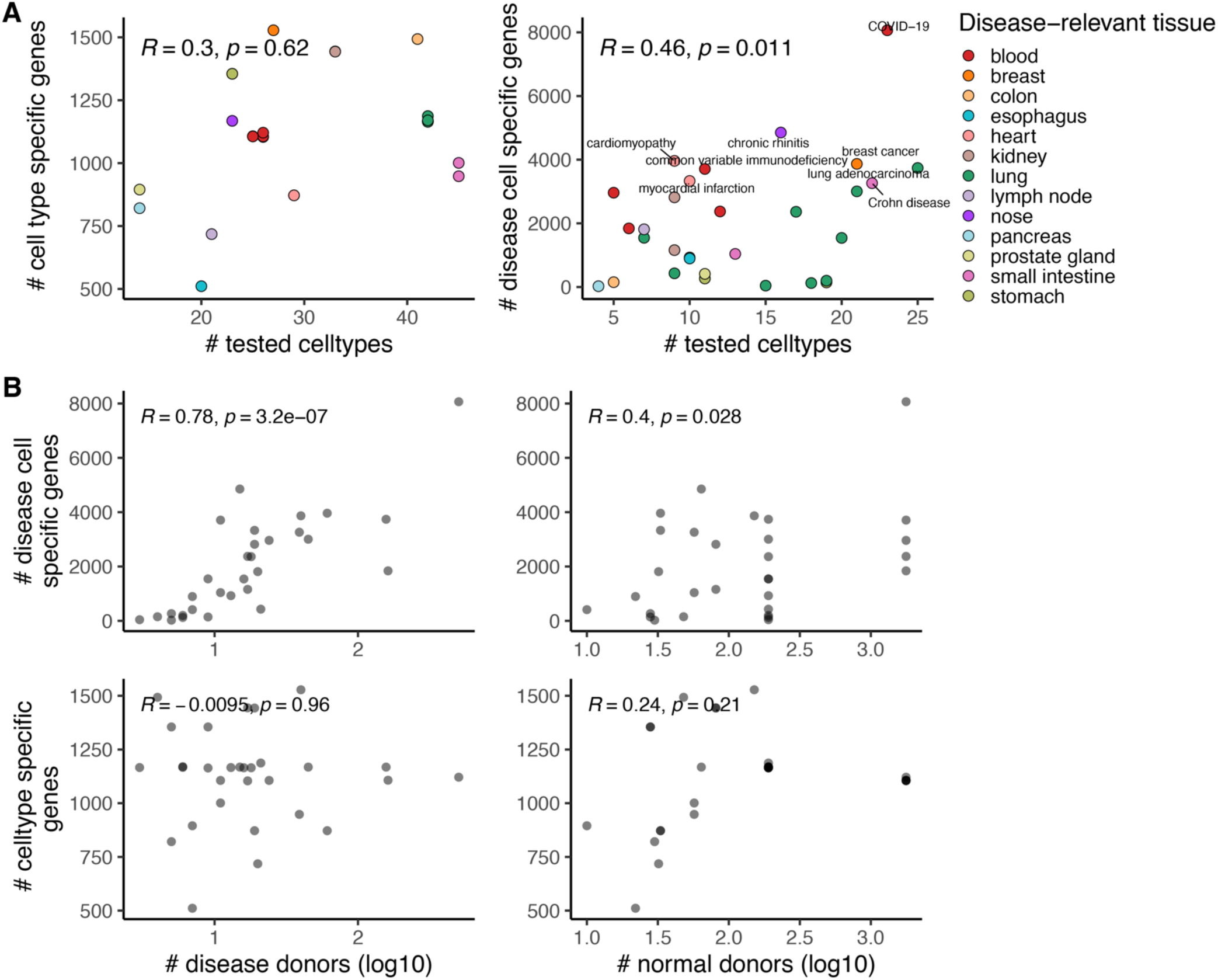
Variability in scRNA-seq supported targets between diseases. (A) Scatterplots showing the number of tested cell types in disease-relevant tissue (x-axis) against the number of identified cell type specific (left) and disease cell specific (right) genes. Dots are colored by disease-relevant tissue. Pearson’s correlation coefficient and p-value for permutation test are shown on top. (B) Scatterplots showing the number of disease donors (left column) and control donors (right column) in scRNA-seq dataset for each disease against the number of identified cell type specific (bottom row) and disease cell specific (top row) genes. Pearson’s correlation coefficient and p-value for permutation test are shown on top. Data is shown for 30 diseases and 13 tissues with data in CellxGene datasets (including diseases that were subsequently excluded because of missing clinical data).

**Supplementary Figure 11:**
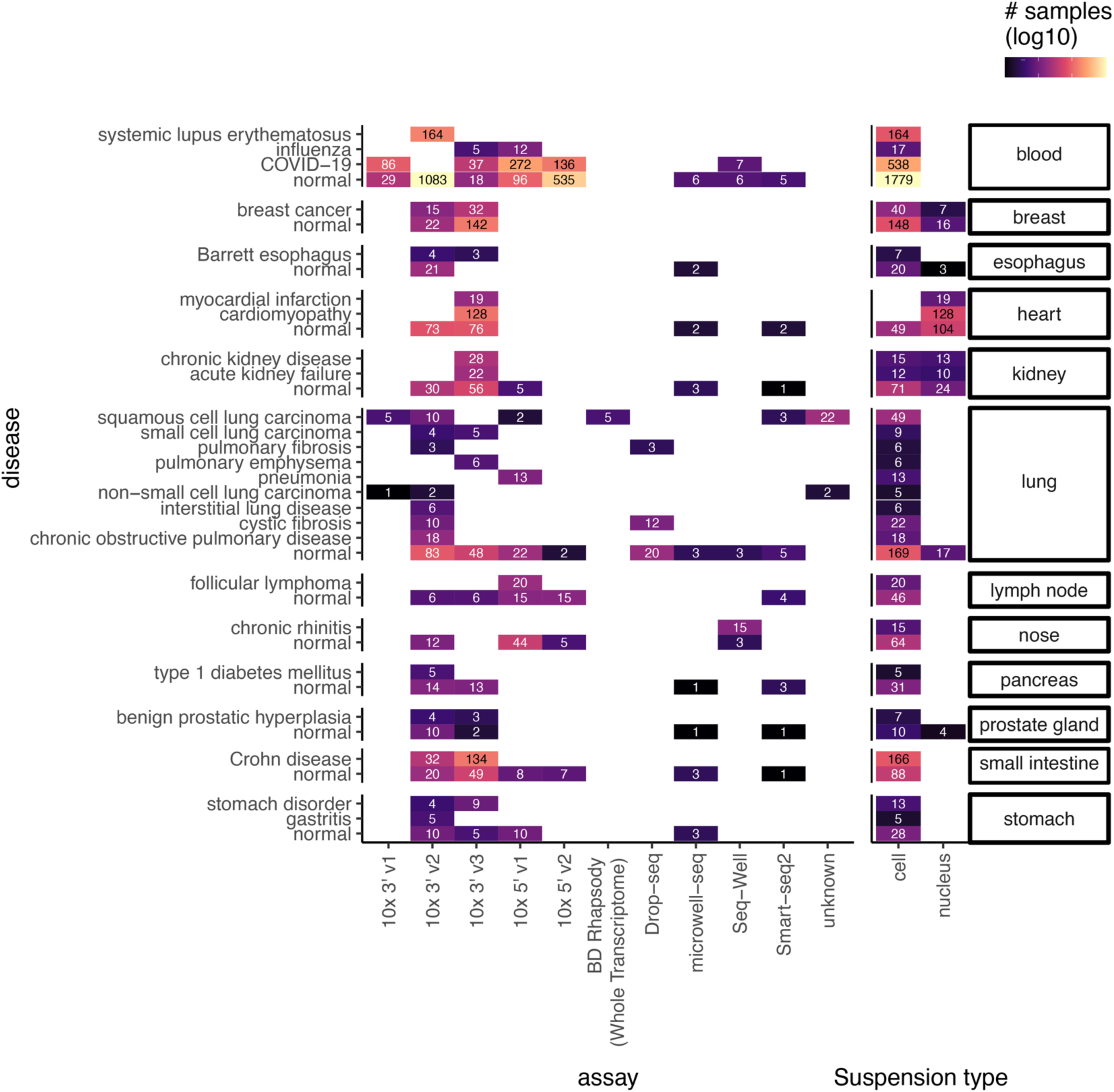
Technical metadata for disease scRNA-seq datasets. Heatmap showing the scRNA-seq assay and suspension type (x-axis) for samples of different tissues and diseases (y-axis). Heatmap color and annotated numbers denote the number of samples analysed for each group. Diseases are grouped by disease-relevant tissue.

## Supplementary Tables

**Supplementary Table 1:** Table of diseases available in CZ CellxGene database considered for study [disease] name of disease used in study

[disease_ontology_id] MONDO identifier for disease used in study

[disease_relevant_tissue] Manually curated annotation for disease-relevant tissue

[disease_name_original] Name of disease found in CZ CellxGene database

[disease_ontology_id _original] MONDO identifier for disease found in CZ CellxGene database

[reason2exclude] if not NA, description of reason to exclude disease from final analysis

**Supplementary Table 2:** Sample-level metadata for scRNA-seq datasets from CZ CellxGene database used in study

[assay] scRNA-seq protocol

[tissue] original tissue annotation

[tissue_general] high-level mapping of a tissue

[suspension type] indicates whether cells or nuclei were isolated

[disease] disease condition of donor

[dataset_id] Identifier for dataset in CellXGene Census

[donor_id] Identifier for donor in dataset

[development_stage_ontology_term_id] Human Developmental Stages ontology term for age of donor

[sample_id] sample identifier (donor, assay, tissue)

[disease_name_original] name of disease found in CZ CellxGene database

[disease_ontology_id _original] MONDO identifier for disease found in CZ CellxGene database

[disease_ontology_id] MONDO identifier for disease used in study

[disease_relevant_tissue] Manually curated annotation for disease-relevant tissue

**Supplementary Table 3:** Results of association analysis between omic support and clinical success across diseases

[odds_ratio] Odds ratio of association between evidence and clinical success

[ci_low] 95% confidence interval of odds ratio (bottom)

[ci_high] 95% confidence interval of odds ratio (top)

[pval] Fisher exact test p-value for enrichment (alternative hypothesis: odds ratio higher than 1)

[n_success] Number of successful gene-disease pairs

[n_insuccess] Number of not successful gene-disease pairs

[n_supported_approved] Number of successful gene-disease pairs supported by omic evidence

[n_supported] Total number of gene-disease pairs supported by omic evidence

[evidence] omic support class (all_sc_evidence indicates cell type and disease cell specific genes)

[clinical status] Clinical success class

[universe] Name of considered gene universe

[universe_size] Number of genes in gene universe

**Supplementary Table 4:** Table of diseases and disease-relevant tissue annotation for expanded cell type specificity analysis

[indication_mesh_term] Name of disease/indication from MeSH

[indication_mesh_id] MeSH ID for disease/indication

[tissue_test] Annotation of disease-relevant tissue used in this study

Supplementary Table 5: Results of association analysis between cell type specificity and clinical success across expanded set of diseases

[odds_ratio] Odds ratio of association between evidence and clinical success

[ci_low] 95% confidence interval of odds ratio (bottom)

[ci_high] 95% confidence interval of odds ratio (top)

[pval] Fisher exact test p-value for enrichment (alternative hypothesis: odds ratio higher than 1)

[n_success] Number of successful gene-disease pairs

[n_insuccess] Number of not successful gene-disease pairs

[n_supported_approved] Number of successful gene-disease pairs supported by omic evidence

[n_supported] Total number of gene-disease pairs supported by omic evidence

[evidence] omic support class

[clinical status] Clinical success class

